# Modeling waning and boosting of COVID-19 in Canada with vaccination

**DOI:** 10.1101/2021.05.18.21257426

**Authors:** Lauren Childs, David W Dick, Zhilan Feng, Jane M Heffernan, Jing Li, Gergely Röst

## Abstract

SARS-CoV-2, the causative agent of COVID-19, has caused devastating health and economic impacts around the globe since its appearance in late 2019. The advent of effective vaccines leads to open questions on how best to vaccinate the population. To address such questions, we developed a model of COVID-19 infection by age that includes the waning and boosting of immunity against SARS-CoV-2 in the context of infection and vaccination. The model also accounts for changes to infectivity of the virus, such as public health mitigation protocols over time, increases in the transmissibility of variants of concern, changes in compliance to mask wearing and social distancing, and changes in testing rates. The model is employed to study public health mitigation and vaccination of the COVID-19 epidemic in Canada, including different vaccination programs (rollout by age), and delays between doses in a two-dose vaccine. We find that the decision to delay the second dose of vaccine is appropriate in the Canadian context. We also find that the benefits of a COVID-19 vaccination program in terms of reductions in infections is increased if vaccination of 15-19 year olds are included in the vaccine rollout.

## Introduction

Several vaccination policy-making bodies, including the Government of Canada’s National Advisory Committee on Immunization (NACI), have utilized transmission modelling studies to evaluate possible strategies for allocating vaccine against SARS-CoV-2. Some policy-making bodies have worked solely with in-house modellers, others with external ones, and still others a combination of both (1; 2), while realizing that models are approximations of natural phenomena, given the quality of available information.

Most contemplated vaccination strategies are based on ethical or practical versus scientific considerations. Determining that healthcare workers (HCWs) should be vaccinated first, followed by other essential workers, institutionalized populations, especially elderly people, and others with chronic conditions that increase their risk of serious illness does not require modelling. Nor does identifying other essential workers. Those strategies involve directly protecting essential or vulnerable groups. Questions amenable to transmission modelling include the rate at which the vaccination program can be expanded to sub-populations not included in the aforementioned list, especially given a limitation in the number of doses available to the population at any given time. Additionally, modelling can assess indirect effects of vaccination. Might, for example, vaccinating some population groups have more impact on transmission than vaccinating others? Such questions are usually framed in terms of population immunity, considering single- and two-dose efficacies of available vaccines (3), as well as the number of doses available to the population. The apparent single-dose efficacy (3), together with our ability to produce and distribute doses, has led some to ask if we should adhere to the two-dose schedule or vaccinate twice as many people once, or vaccinate twice as many people once as soon as possible. Similarly, while the currently available vaccines seem efficacious in the short term for the age groups tested in the clinical trials, we have no idea how long such protection will last. We thus must consider the indirect effects of protecting certain population cohorts through vaccination in others, not just their caregivers, but other members of the general population as well.

Most models of the transmission of SARS-CoV-2 are modifications of a classic model in population biology in which the host population is partitioned into those who are susceptible to infection; infected, but not yet infectious; infectious; and removed from the process (e.g., (4–7)). These modifications include features of the biology of COVID-19 that might affect transmission such as pre- and asymptomatic infections, hospitalization of some with symptomatic infections, vaccination, mortality, and waning of immunity (e.g., (8–16)). To answer some of these questions in the context of pertussis (prior to the SARS-CoV-2 outbreak), we used a model with some of these features that emphasizes the waning and boosting of immunity and, furthermore, the relationship between immunity when infected and disease (17). We extend our model structure here to COVID-19.

We believe that economic calculations should be based on sound mathematical epidemiology, which we strove to provide, but are not economists, so have left economic questions for those with the requisite expertise. We also endeavored to make the case for vaccinating adolescents, relatively few of whom experience serious disease, but who – by virtue of their contacts – can be super-spreaders. Consequently, our answer to the question, “Might vaccinating some groups have more impact on transmission than vaccinating others?” is “Yes.” The answer might differ for other policy goals (e.g., reducing deaths).

## Model

We have implemented a model of COVID-19 infection with age structure (i.e., groups 0-4, 5-9, …, 75+ years). A flow diagram of the model is shown in Figure 1 for a single age group. The model is based on a Susceptible-Exposed-Infected-Vaccinated-Susceptible model structure (SEIVS). We use *S*_*i*_, 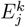, *I*_*j*_, and 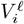 to denote the number of susceptible, exposed, infectious and vaccinated individuals in each age group, where *i* (1≤ *I* ≤ 4) denotes immune status, *j* (1 ≤ *j* ≤ 3) denotes symptom severity, *k* (1 ≤ *k* ≤ 3) represents stages in the exposed class (to obtain gamma-distributed exposed sojourns), and *ℓ* = 1, 2 denotes the number of doses of vaccine that individuals have received.

**Figure 1:**
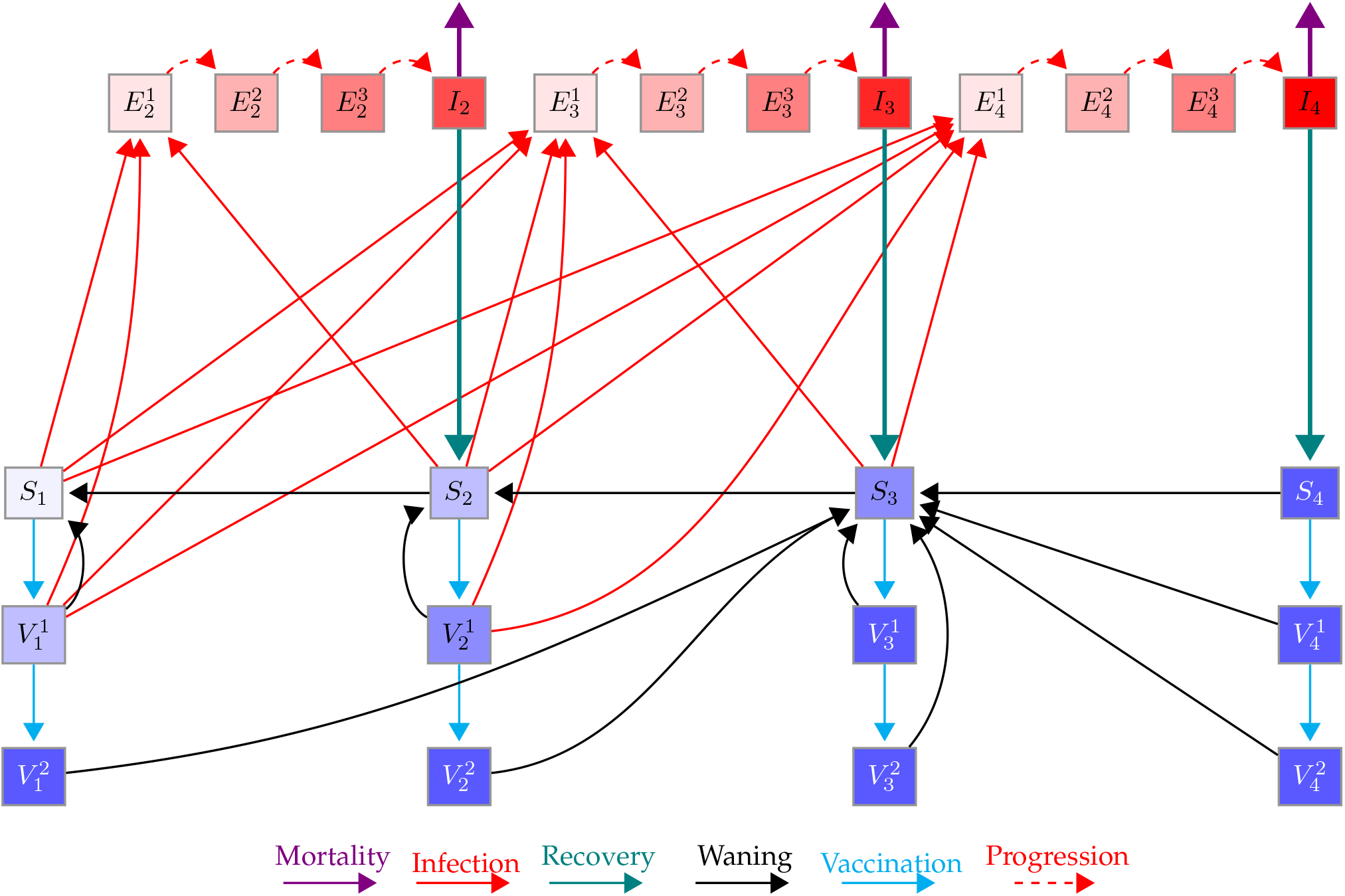
Schematic of the model for one age group. Here, *S*_1_, *S*_2_, *S*_3_, and *S*_4_ (purple shaded boxes) represent susceptible individuals who are immunologically naive, have some, moderate, and full immunity, respectively. *I*_2_, *I*_3_, and *I*_4_ (red boxes) represent infected individuals with mild, moderate and severe symptoms, respectively, who will develop some, moderate, and full immunity once recovered (teal lines), respectively. 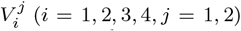 (*i* = 1, 2, 3, 4, *j* = 1, 2) represent vaccinated individuals from the *S*_*i*_ classes (*i* = 1, 2, 3, 4) after *j* = 1, 2 doses of vaccine given a two-dose schedule. 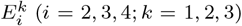(*i* = 2, 3, 4; *k* = 1, 2, 3) represent exposed individuals (infected, asymptomatic, not infectious) with progressive stages *k* = 1, 2, 3 that will experience mild *I*_2_, moderate *I*_3_, and severe *I*_4_ symptoms. Susceptible and vaccinated individuals can be infected and move to the exposed classes (red lines). Susceptible and vaccinated classes at the same location on the immunity continuum have similar characteristics. Immunity gained from infection and vaccination can wane (black lines).

The base model consists of an immune continuum along which we distinguish four states (fully susceptible (*S*_1_), somewhat immune (*S*_2_), moderately immune (*S*_3_), and fully resistant to infection (*S*_4_)), three infectious states with mild (*I*_2_), moderate (*I*_3_), and severe (*I*_4_) symptoms, and three infected but non-yet-infectious states (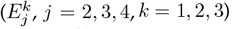, *j* = 2, 3, 4, *k* = 1, 2, 3). We assume that individuals of higher immune status are less susceptible to infection than those of lower status. As per (18), we assume that co-morbidity determines the probability of mild, moderate, and severe symptoms for each age group. Immunity develops after infection, and we assume that people with mild, moderate, and severe symptoms move to immune classes *S*_2_, *S*_3_ and *S*_4_, respectively, upon recovery. This is tantamount to assuming that severity of symptoms is proportional to neutralizing immunity development (19–21). Finally, we assume that immunity wanes over time (Figure 1, black lines), a characteristic common to other known coronaviruses (22).

We extend the base model to include two vaccinated states per immune state *S*_*i*_, corresponding to one 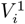 and two 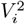 vaccine doses. We distinguish the susceptible and vaccinated states solely to facilitate calculating coverage. Here, we assume that two doses of vaccine provide the same level of immunity as *S*_4_; i.e., people in 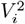 have similar characteristics to those in *S*_4_. Additionally, we assume that one dose of vaccine administered to individuals in immune states *S*_3_ and *S*_4_ also results in resistance to infection; i.e., 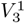 and 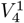 have similar characteristics to *S*_4_. Finally, we assume that one dose of vaccine given to individuals in *S*_1_ and *S*_2_ provides immunity similar to states *S*_2_ and *S*_3_, respectively. In other words, 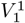 has similar characteristics to *S*_2_ and 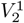 to *S*_3_. Concurrently, we assume that immunity can wane from the vaccinated classes as from their corresponding *S* classes (Figure 1, black lines).

A detailed model description is provided in the Supplementary Material, including parameter values and references. Briefly, we track mild, moderate and severe symptoms by age given age-specific model parameters defining the population contact structure during successive public health mitigation phases throughout the COVID-19 pandemic. We then add a vaccination program and track vaccine doses by age under specific scenarios of interest. We quantify vaccination program outcomes by calculating the number of infections averted and percent reduction in the number of infections with mild, moderate, and severe symptoms by age. We consider parameters specific to the Canadian population and public health mitigation strategies, including variations in contacts between age groups, in school, work and other settings, to reflect school closures, social distancing measures and work-from-home orders. We further reduce the contact matrix by a factor reflecting the use of personal protective equipment (PPE), hand-washing, and other non-pharmaceutical interventions (NPIs).

We consider vaccination programs as described by current vaccination coverage data (23) and the National Advisory Committee on Immunization vaccination program description (24). We also consider vaccination programs including the prioritization of certain age groups and other sub-populations (e.g., HCWs). To quantify the outcomes of the current vaccination program under way in Canada (a two-dose program with 16 weeks separating the first and second doses), we compare this program to one-dose scenarios, including a one-dose vaccine that provides neutralizing immunity (vaccination moves individuals to 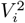 classes whereby they are resistant to infection), a one-dose vaccine that provides some non-neutralizing immunity (vaccination moves individuals to 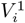 classes, whereby those in classes 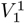 and 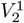 have some susceptibility to infection), and a two-dose vaccine program that covers only half of the target population, but separates the doses using the vaccine-developed-recommended interval of 28 days. In the two-dose program, individuals move to 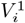 classes first, and then to 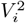 classes after the prescribed period (28 days or 16 weeks). We note that, to compare one-versus two-dose programs with the same amount of vaccine (i.e., when supply is limited), it is sufficient to compare one-dose delivery with full target coverage to two-dose delivery with half target coverage. We also note that results of a program with a delayed (beyond the interval recommended by the manufacturer, or the government chosen time frame) second dose will lie between those of the one-dose program with non-neutralizing immunity and the two-dose program with full target coverage with no delay between doses (not considered here). The vaccination programs are described in Table 1.

**Table 1:** Vaccination Programs. Scenario 1, under two-dose coverage with 16 weeks between doses has been designed to resemble the actual COVID-19 vaccination program in Canada from January to April 2020. *Assume that HCWs are aged 20-64 years and evenly distributed over each 5-year age group in this interval.

|  |  |  |
| --- | --- | --- |
| Dosage | One-dose: | Full target coverage<br>With neutralizing immunity moves individuals to $V_i^2$. Without neutralizing immunity moves individuals to $V_i^1$. |
| | Two doses: | [half, full] target coverage<br>Doses separated by 4 weeks for half coverage, and 16 weeks for full coverage.<br>First dose moves individuals to $V_i^1$. Second doses moves individuals to $V_i^2$. |
| Coverage | Target: | 80% of the Canadian population |
|  | Monthly coverage: | 2.4%, 2.7%, 8.9%, 19.0%, 18.0%, 18.0%, 10.2%, 1.0% |
| Scenario |  |  |
| 1 |  | Vaccinate healthcare workers (HCWs)* (months 1-2)<br>Vaccinate ages 65+ years (months 1-8)<br>Vaccinate ages 60-64 years (months 3-8)<br>Vaccinate ages 50-59 years (months 4-8)<br>Vaccinate ages 45-49 years (months 5-8)<br>Vaccinate ages 20-44 years (months 6-8) |
| 2 |  | Vaccinate healthcare workers (HCWs)* (months 1-2)<br>Vaccinate ages 65+ years (months 1-8)<br>Vaccinate ages 55-64 years (months 3-8)<br>Vaccinate ages 50-54 years (months 4-8)<br>Vaccinate ages 45-49 years (months 5-8)<br>Vaccinate ages 20-44 years (months 6-8) |
| 3 |  | Vaccinate healthcare workers (HCWs)* (months 1-2)<br>Vaccinate ages 65+ years (months 1-8)<br>Vaccinate ages 50-64 years (months 3-8)<br>Vaccinate ages 40-49 years (months 4-8)<br>Vaccinate ages 25-39 years (months 5-8)<br>Vaccinate ages 20-24 years (months 6-8) |
| 4 |  | Vaccinate all adults 20+ years of age (months 1-8) |

Vaccination programs are implemented on a week-by-week basis until a target coverage level is achieved. The target coverage level takes into account the age groups that have been approved to receive a COVID-19 vaccine (see Table 1). Given the desired coverage, we determine the requisite vaccination rate (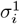, where *i* represents the immune state) for each age group. To model a two-dose program, we determine the rate for the first dose as for a single dose and the rate for second dose given the recommended interval between doses (provided by the vaccine manufacturer, or the government chosen time frame). We note that, when we calculate vaccination rates for the first dose, we remove individuals who were already vaccinated or have known infection (e.g., *I*_4_). Consequently, we allow people unaware that they have been or are currently infected to be vaccinated; for example, those in the exposed classes, or who are asymptomatic or have mild symptoms (some fraction of the mild and moderate states). In these cases, we assume that vaccination does not alter the level of immunity acquired; that is, we assume that ensuing immunity depends solely on the severity of symptoms currently experienced (i.e., those vaccinated from 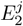,*I*_2_ and 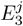,*I*_3_ follow their disease course and recover to *S*_2_ and *S*_3_, respectively). This is a reasonable assumption insofar that an infected individual’s immune system would be primed at the time of vaccination.

While HCWs represent priority groups for vaccination, we do not explicitly model these sub-populations. Age distribution information for these groups, however, is available (25–28). Herewith, we assume a uniform distribution of HCWs over each 5-year age group from 20 to 64 years (see Supplementary Material for details).

The model incorporates public health mitigation strategies as adapted by the Canadian population, on average (since mitigation can vary between different jurisdictions). Specific modifications to the contact matrices are provided in the Supplementary Material. In the past, the choice of mitigation matrix is informed by the mitigation strategies adapted by different provinces and territories. However, as we cannot predict the future, we must adapt specific contact matrix structures projecting forward in time. Given the time frame under consideration, Summer and Fall 2021, we have chosen to adopt the same matrices implemented over Summer and early Fall 2020.

### Temporal Changes in Testing Rate and Contact Tracing

While temporal changes in contact structure, as affected by different public health mitigation strategies over time, and temporal changes in vaccine availability to different age groups have been addressed explicitly in our modelling structure, using modified contact matrices and calculation of different vaccination rates (see above), we have not explicitly incorporated changes in the testing rate, or in contact tracing activities. Both these, however, affect the transmission of the virus. We therefore assume that temporal variation in testing and contact tracing can be captured in the parameter *κ* in our model, which also is used to reflect variation in PPE and social distancing compliance. Parameter *κ* is the only model parameter that is fit to COVID-19 data; therefore, it can capture changes in behaviour and pubic health mitigation over time.

### Variants of Concern

In recent months, different variants of the SARS-COV-2 virus have emerged (29). It has been reported that these variants may be up to 1.5 times more transmissible than the original wild type strain (30–34). Our mathematical model can incorporate the difference in transmission, again, using parameter *κ*, the only model parameter that is fit to COVID-19 data. As transmissibility increases in the population, the value of *κ* will naturally increase to reflect this effect in the COVID-19 data.

It is believed that the effectiveness of currently approved vaccines will be minimally altered by the variants of concern (35–37). We therefore choose to keep parameters reflecting vaccine efficacy to be constant even when the variants of concern exist in the circulation of the virus in the population.

## Results

### Model Fitting

Figure 2 plots the model fit from January 2020 to April 15, 2021. The daily reported infections (crosses) from (23) and simulated daily incidence of *I*_4_ severe (solid line), *I*_3_ + *I*_4_ moderate + severe (dotted line), and *I*_2_ + *I*_3_ + *I*_4_ mild + moderate + severe (dashed line) infections for the entire population are shown. The model is fit using parameter *κ*, which reflects the population compliance to public health mitigation factors, and incorporates temporal variations in testing, contact tracing and transmissibility of the virus (as variants of concern infiltrate the population). Parameter *κ* is determined so that daily reported incidence lies between the severe (solid line) and moderate + severe (dotted line) curves. The model fitting also incorporates the different public health mitigation periods of various strengths, that are reflected using modified contact matrices for work-from-home, school closure, business closures, etc (see Table S4), and the Canadian vaccine roll-out program (see Table 1) starting on January 15, 2021. Additionally, parameters were determined so that the basic reproduction number *R*_0_ corresponds to the median value reported for the Canadian COVID-19 epidemic, *R*_0_ = 2.6 (18). Given the correspondence illustrated, we plot age-specific predictions in Supplementary Figure S1. Additionally, in the Supplementary Figure S2, we plot the reduction in the average number of contacts for the entire population over time, given modifications in the contact matrices (x) and the combined effect of the modifications in the contact matrices and the fitted value of parameter *κ* (+).

**Figure 2:**
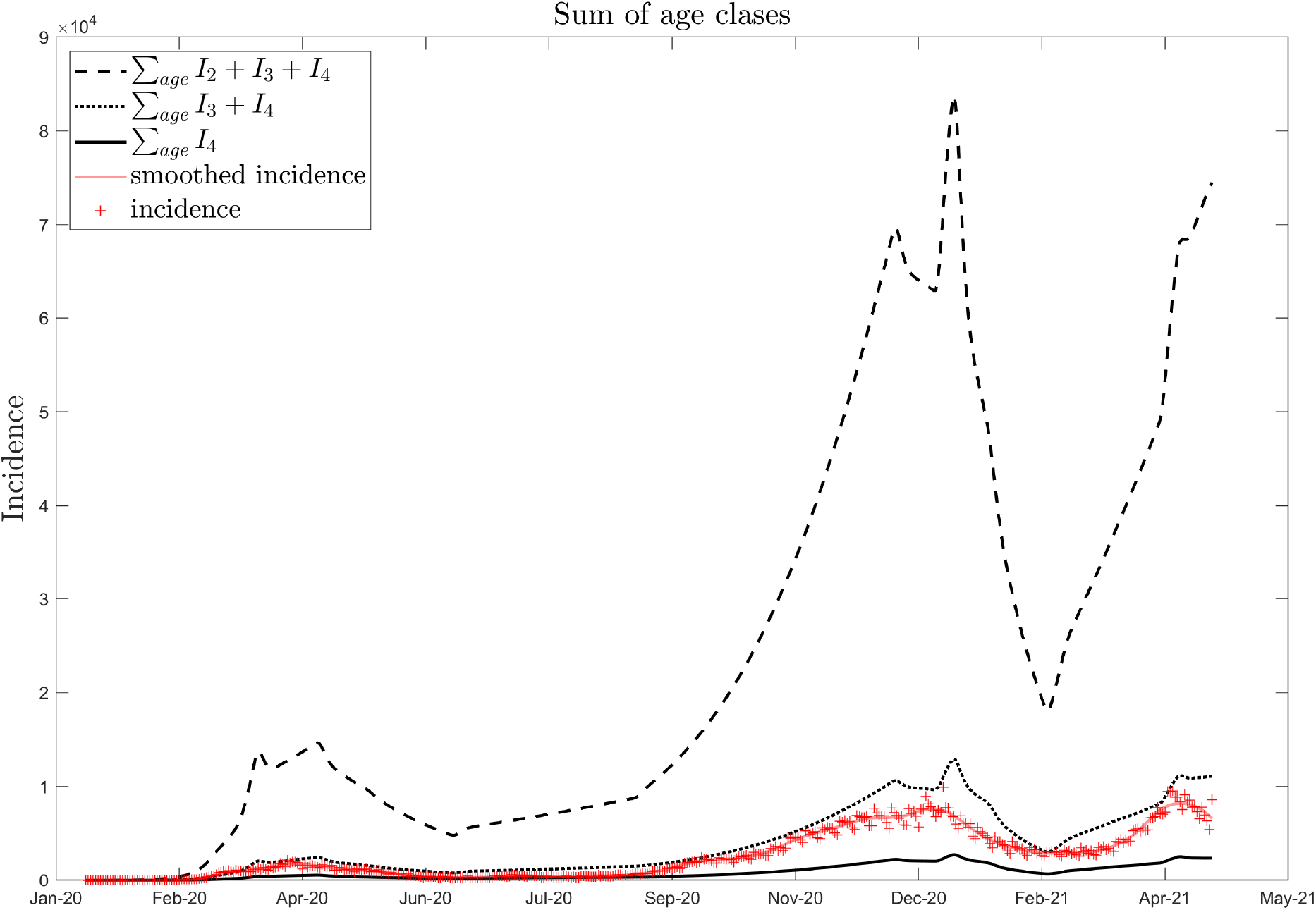
Daily incidence. Predicted incidence of severe (*I*_4_, solid line), moderate + severe infections (*I*_3_ + *I*_4_, dotted line), and mild + moderate + severe infections (*I*_2_ + *I*_3_ + *I*_4_, dashed line) are shown. Daily COVID-19 reports from (23) are shown (red crosses), and loess smoothed in Matlab (red line).

### Vaccination

Vaccination, no matter the type of one- or two-dose vaccine considered here (see Table 1) always reduces the number of infections in every age group (Figures 3 and S3). Intuitively, a one-dose neutralizing vaccine (blue lines) has the best outcome. A two-dose vaccine at half target coverage with 28 days between doses (black lines) performs the worst in the short-term, and a one-dose non-neutralizing vaccine performs the worst in the long-term. For all one- and two-dose vaccines, the best outcome for the entire population is realized when all adults 20+ years of age are vaccinated over the entire vaccination program (Scenario 4, dotted lines), except when considering a one-dose non-neutralizing vaccine, when the Scenario 1 vaccine program slightly outperforms all others.

**Figure 3:**
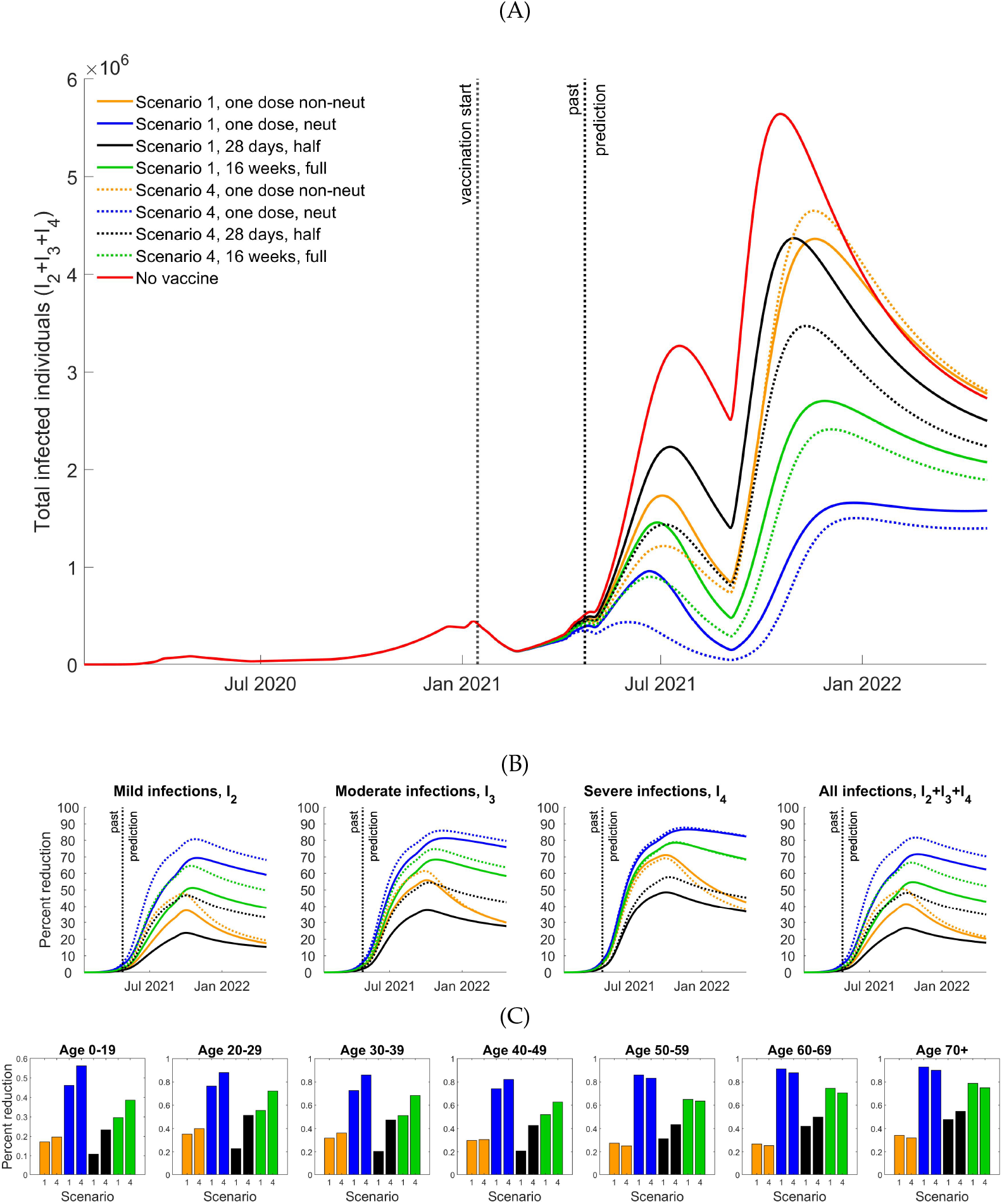
Effect of vaccination, Scenarios 1 and 4. (A) Infected populations without (red line) and with one dose that is non-neutralizing (blue), one dose that is neutralizing (orange), two doses at half target coverage with 28 days between doses (black) and two doses at full target coverage with 16 weeks between doses (green). (B) Percent reduction in *I*_2_, *I*_3_, *I*_4_, and *I*_2_ + *I*_3_ + *I*_4_ infections over time. (C) Percent reduction on day 365 for each age group given one-dose non-neutralizing (orange), one-dose neutralizing (blue), two-dose half target coverage (black) and two-dose full target coverage (green) vaccines.

To compare the epidemic outcome with no vaccine (red line) to each vaccination program, we plot the percent reduction in the number of *I*_2_, *I*_3_, *I*_4_, and *I*_2_ + *I*_3_ + *I*_4_ cases over time, projected for a year after initiation of the vaccination program. These are plotted in Figure 3 panels (B), and for specified age groups, in panel (C). Panel (B) shows that the best overall reduction in all infections (right) occurs under Scenario 4, when adults aged 20+ years are vaccinated every month. In panel (C), however, we observe that reductions in severe infections in age groups 50+ years are greatest under Scenario 1, except for the two-dose vaccine with half coverage and 28 days between doses (black). We only present results for Scenarios 1 and 4 in the main text, but results for Scenarios 2 and 3 are provided in the Supplementary Material. In addition, Figure 3 shows that the relative ordering of effectiveness of strategies is consistent over time, despite differing by the specific group considered, e.g. young versus older adults. This indicates that measurements of reduction at any time point should give a comparative ordering of strategies.

The *I*_4_, and total (*I*_2_ + *I*_3_ + *I*_4_) infections averted given a two-dose, full target coverage vaccination program (with 16 weeks between doses) under Scenarios 1 and 4 are shown in Figure 4 (see Supplementary Figure S4 for all scenarios under a two-dose, full target coverage, vaccination program). We observe that the number of *I*_4_ and total infections averted due to vaccination continue to increase beyond the vaccination program (July 2021). Once again, it is also evident that Scenario 4 results in the maximum total cases averted, but Scenario 1 results in the greatest number averted among those aged 60+ years (see Figure 4, last column).

**Figure 4:**
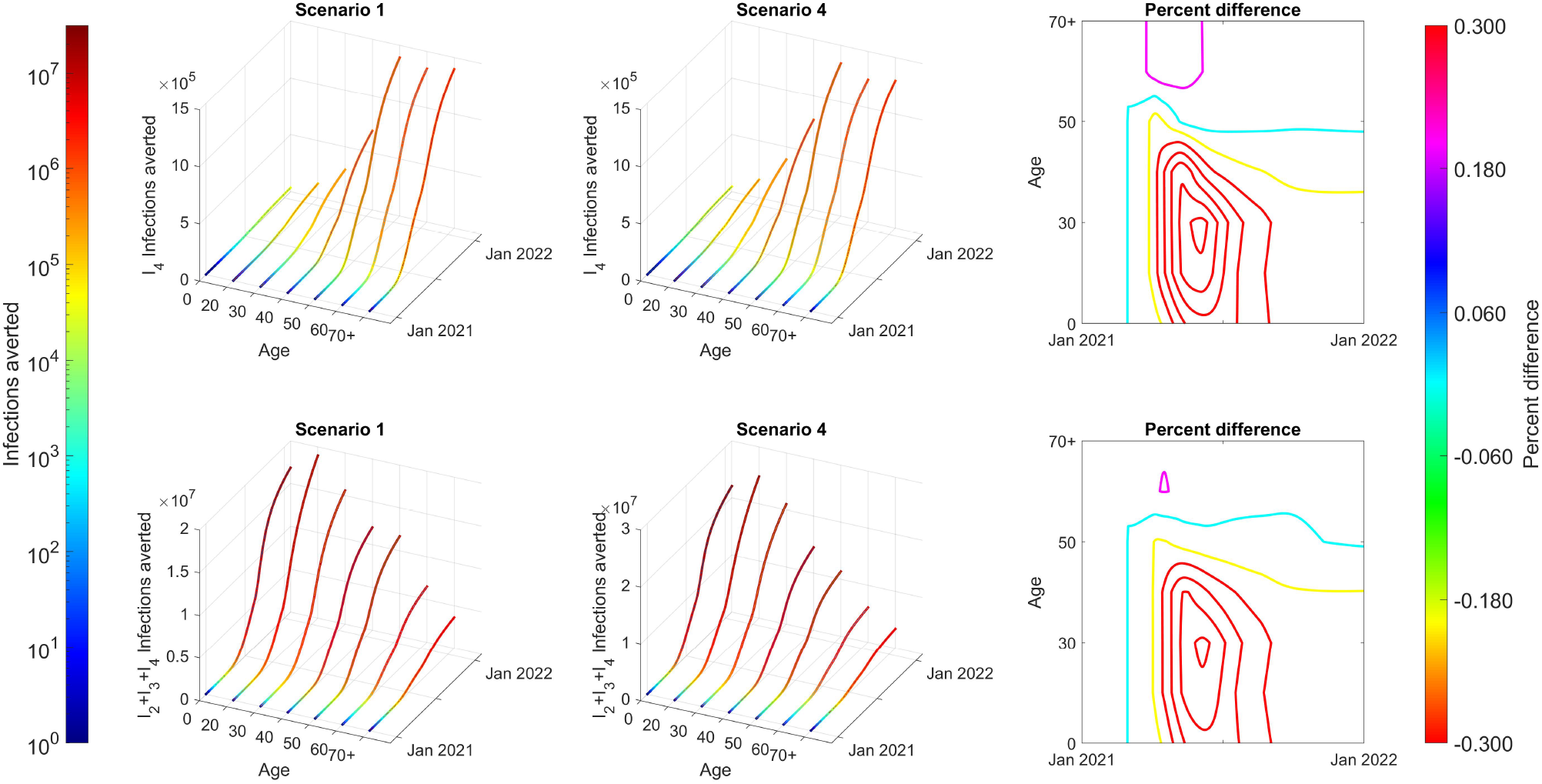
Infections averted: Scenarios 1 and 4, two-dose, full target coverage, with 16 weeks between doses. Infections averted in *I*_4_ (top row) and *I*_2_ + *I*_3_ + *I*_4_ (bottom row) given Scenarios 1 (left column) and 4 (middle column). The percent difference between Scenarios 1 and 4, calculated as ([Scenario 1 infections averted] - [Scenario 4 infections averted])/[Scenario 1 infections averted]) is shown (last column). Here, infections averted are determined by the difference between cumulative incidence with no vaccine and under the vaccination program considered).

Vaccination programs targeting specific age groups will change the age distribution of new infections. In turn, changes in the age distribution of reported cases will occur. It is therefore important to determine what changes in the age distribution should be expected, so that such changes are not misinterpreted as vaccination program failure, or shifts in infection target by the pathogen. Figure 5 plots the age distribution of daily *I*_4_ (left column) and *I*_2_ + *I*_3_ + *I*_4_ infections (right column) for Scenarios 1 (panel A) and 4 (panel B) given a two-dose, full target coverage vaccination program with 16 weeks between doses. We see that the age distribution of *I*_4_ and *I*_2_ + *I*_3_ + *I*_4_ infections varies for both scenarios during and after the vaccination program, including a shift to a higher fraction of severe cases reported in younger age groups. This, however, does not mean that there is a shift of COVID-19 targets of infection. This merely reflects the distribution of the vaccine to older ages. Supplementary Figure S5 provides the same information for the other vaccines and vaccination programs listed in Table 1.

**Figure 5:**
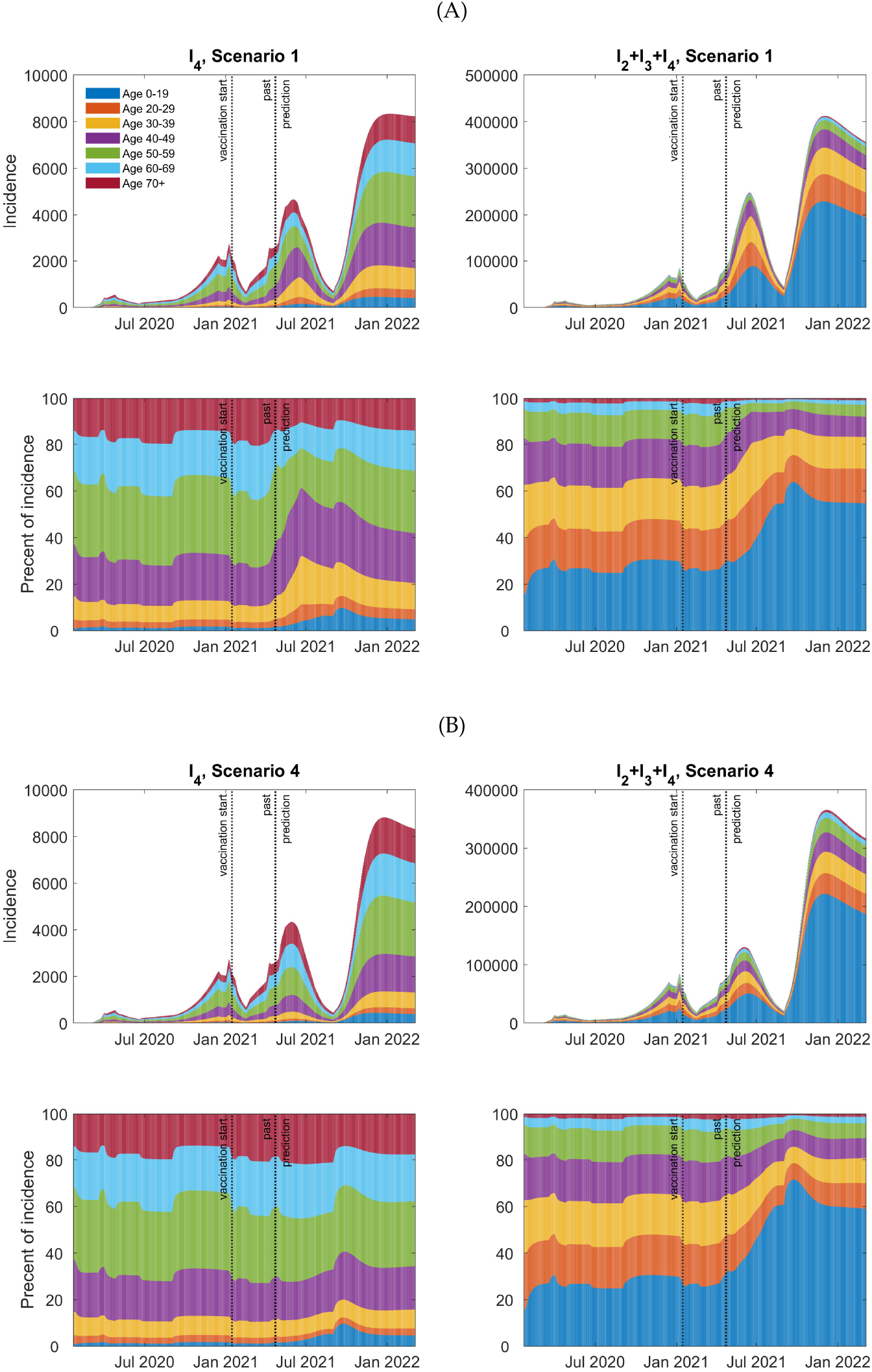
Age distribution of daily incidence: two-dose, full target coverage with 16 weeks between doses. (panel A) Scenario 1. (panel B) Scenario 4. In both panels, we plot the daily incidence of *I*_4_ and *I*_2_ + *I*_3_ + *I*_4_ infections by age (top row), and by the percent in each group (bottom row).

#### Indirect Vaccine Effects

Vaccination not only protects vaccinated individuals, it also protects others. In Figure 6, we plot the percent reduction in *I*_4_ infections during the year after initiation of a vaccine program assuming that only one group is vaccinated at a time. To quantify the indirect effect, we vaccinate individuals in a single age group to a maximum 0.5% of the Canadian population each month for six months. That is, we determine the fraction of the population in each age group that needs to be vaccinated so that 0.5% of the entire Canadian population is vaccinated in one month, and up to 3% over 6 months. Figure 6 (left panel, and Supplementary Figure S6, which plots the percent reduction in *I*_2_, *I*_3_ and *I*_4_ infections 90, 180 and 365 days after vaccination program initiation) clearly shows the direct effect of vaccination within a single age group (the diagonal elements). We also observe that vaccination of any age group benefits others (off-diagonal elements). These indirect effects are most pronounced for the 15-19 year age group. Vaccination coverage of 3% of this population reduces infections in other age groups by approximately 10% (light blue shading) during the year that vaccination begins.

**Figure 6:**
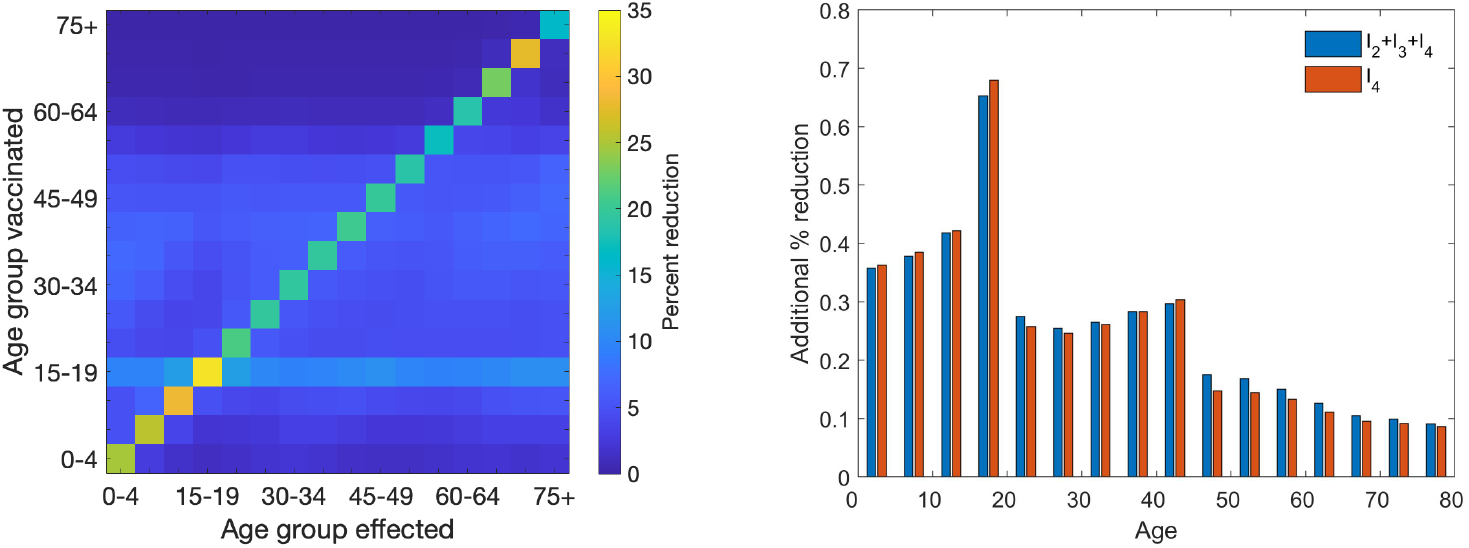
Indirect effects of vaccination. (left) The percent reduction (colour) in the number of severe infections for each age group (x-axis), given two-dose vaccination with 28 days separating doses, and two weeks until full realization of neutralizing immunity) of one age group (y-axis). Vaccination rates are calculated so that roughly 0.5% of the Canadian population is vaccinated each month for six months (up to 3% coverage over 6 months) whereby all vaccinated individuals are taken from a single age group (y-axis). (right) The further reduction in infections of *I*_2_ + *I*_3_ + *I*_4_ (blue bars) and *I*_4_ (red bars) when including vaccination of 16-19 year-olds in the final two months of a two-dose full target coverage vaccination program with 16 weeks between doses.

Until very recently, COVID-19 vaccines in Canada were not approved for use in children under the age of 16 (38). To maximize benefit of a vaccination program in the younger age groups, vaccination of 16-19 year olds (in age group 15-19) should be considered. It is important to ensure, however, that vaccination of 16-19 year olds would not decrease the benefit of vaccination among the elderly. To quantify the benefit of vaccinating 16-19 year olds, we added this age group to those to which vaccines are distributed in the final two months of the vaccination programs (see Table 1). Overall, we find that redistribution to include vaccination of 16-19 year-olds benefits the entire population. All age groups experience increments in percent reduction of COVID-19, from 10 to 65 percentage points (Figure 6, right panel).

#### Restrictions in Vaccine Supply

Given restrictions in supply, and wanting to maximize the benefit of a vaccination program, it is necessary to quantify differences in outcome of programs that administer one dose to twice as many people as could be fully vaccinated (with 2 doses each). We now compare and contrast the two-dose, half target coverage (doses 28 days apart), and one-dose non-neutralizing programs studied here. Figure 3 demonstrates that under the vaccine parameters assumed, which are representative of the Moderna, Pfizer and AstraZeneca vaccines, a one-dose non-neutralizing vaccine (orange) will outperform a two-dose vaccine half-coverage (black) program in the short-term, and even some months after the vaccination program has ended (July 2021). We observe, thus, that given anticipation of limitations in vaccine procurement (i.e., shipment or production delays) to accomplish a two-dose full target coverage vaccination program with 28 days between doses, it seems appropriate to administer one dose of vaccine, and allow for a delay in the second dose until shipments can be procured. Early into the vaccination program, Canadian officials opted to modify the program to allow for a delayed second dose. These results justify this early choice.

#### COVID-19 Resurgence in Fall 2021

Figures 3 and 5 (and Supplementary Figures S3 and S5) show that a resurgence of COVID-19 infection may occur in Fall 2021. We note that the magnitude of a resurgence will depend on many parameters, including, vaccine uptake (in all age groups), public health mitigation and relaxation during and beyond Summer 2021, population compliance to PPE wearing and social distancing, testing rates, contact tracing levels, the transmissibility of the prevalent variant strains at the time, and the waning rate of immunity. Therefore, the forecasting for Fall 2021 is very complex. We leave such considerations for future work.

## Discussion

We developed a model of COVID-19 infection by age that includes the waning and boosting of immunity against SARS-CoV-2 due to infection or vaccination. The model, which is first evaluated over different mitigation and vaccination phases (from January 2020 to April 2021) of the COVID-19 epidemic in Canada, is used to study various COVID-19 vaccination programs (24), including administration of one and two doses and different scenarios that involve prioritization of certain groups. As per current public health programming, we implement the vaccination programs using the current assumed mitigation matrix Phase 3 that involves moderate relaxation of restrictions on school, work and other contacts (see Supplementary Material, Demographic Parameters), but additional changes in contact transmission using a linear factor reflecting the use of masks and other PPE, hand-washing, testing rate, contact tracing, and the increased transmissibility of variants of concern. Intuitively, we find that a one-dose neutralizing vaccine has the best outcome under each coverage program, Scenarios 1-4. Model results, however, can also be used to assess outcomes of programs that (1) provide one dose to twice as many people versus two doses to half as many, and (2) delays in providing the second dose of two-dose vaccines. Ultimately, given the parameters assumed here, representative of two-dose vaccines currently approved for use in Canada, we find that, if needed, delivery of one dose to a larger number of people can provide a slightly better outcome than delivering a two-dose program to half as many people, at least into the Fall of 2021. However, we recommend that delays in delivery of a second dose, when it comes available, should be minimized to realize the maximum benefit from a two-dose vaccination program. Ultimately, we find that the early decision of the Canadian government to allow 16 weeks between vaccine doses will achieve better outcomes than a one-dose non-neutralizing scenario, and a two-dose half coverage scenario with 28 days between vaccine doses.

During a vaccination program, compliance with physical/social distancing, use of masks and other PPE and other NPIs may wane. In current work, we are developing relaxation scenarios that will allow (a) certain economic sectors to be re-opened in stages, and (b) some relaxation in public health programming or PPE and/or other NPI compliance. Preliminary simulations show that mild relaxation too early can erode the gains from a vaccination program. Additionally, the existence and magnitude of a Fall 2021 resurgence depends on these factors. A study of relaxation and COVID-19 resurgence remains a course for immediate work.

Some models of the COVID-19 pandemic have incorporated the effects of waning immunity (11–14). To our knowledge, no other model of COVID-19 infection or vaccination considers differential immunity development after infection or vaccination with waning immunity and age structure. Our model can provide distributions of immunity by age before, during, and after a vaccination program. An added benefit thus lies in the utility of our model structure to permit studying vaccination programming that maximizes immunity generation and boosting in the population while minimizing severe COVID-19 illnesses and deaths. Additionally, it allows for better-informed estimates of the reproduction number. Studies of optimal vaccination programs and immunity distribution-informed reproduction numbers are planned.

## Data Availability

All data referred to in the manuscript is publicly available with the reference provided in the manuscript.

## Acknowledgments

We especially thank Dr John Glasser (Centers for Disease Control and Prevention) for assistance and discussion. The findings and conclusions in this report are those of the authors and do not necessarily represent the official positions of the Centers for Disease Control and Prevention, National Science Foundation, or other institutions with which the authors are affiliated. This work was funded by the Natural Science and Engineering Research Council of Canada (JMH,DD), the CIHR-Fields COVID-19 Task Force (JMH,DD), NSF Grant # 2029262 (LMC), Hungarian Scientific Research Fund NKFIH FK 124016 (GR), NSF Grant DMS-1814545 (ZF), NSF IR/D program (ZF), and CSUN 2021-22 RSCA Awards (JL). The authors acknowledge the support of the American Institute of Mathematics (AIM) via workshop and SQuaRE grants.

## Supplementary Material

### Supplementary Figures

#### Infection Incidence by Age as per Model Fitting Results

Figure S1 compares model predictions to reported incidence for each age group. This figure shows that, given model assumptions, incidence by age may lie closer to the severe *I*_4_ infection incidence (solid line), moderate + severe *I*_3_ +*I*_4_ infection incidence (dotted line), and mild + moderate + severe *I*_2_ + *I*_3_ + *I*_4_ incidence (dashed lines). We note that, because the model does not incorporate spatial clusters, there are some instances where reported infections lie very close to, or even greater than, the mild + moderate + severe *I*_2_ + *I*_3_ + *I*_4_ incidence (dashed line). However, the model and reported data are still in close agreement. We note that, when reported infections lie close to the dotted line, this may reflect increased testing in particular age groups; i.e., for younger ages, when school is open, and the elderly in long-term care facilities. We also note that model predictions suggest that adults aged 50-69 do not get tested unless they are symptomatic.

There is an increase in the moderate and mild cases in elderly populations in the early months of 2021. Such increases reflect the increased transmissibility of the variants of concern and the fact that individuals that have experienced waning immunity after infection can experience mild or moderate disease upon reinfection. However, the magnitude of incidence of the moderate and mild infections depends on the assumed rate of waning immunity. A sensitivity analysis on the waning rate is planned for future work.

**Figure S1:**
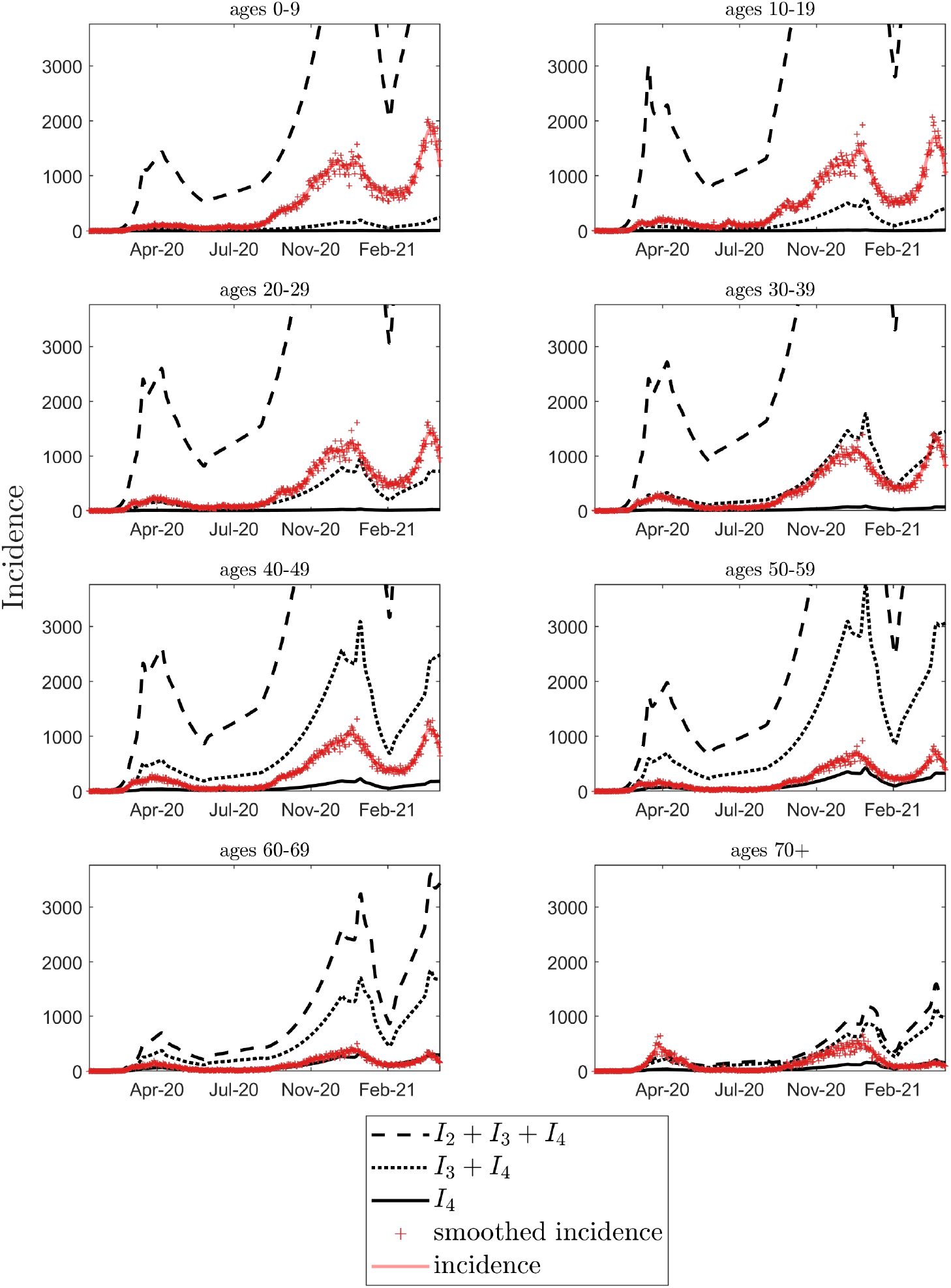
Daily incidence by age, from January 2020 to April 15, 2021. Daily incidence of severe (solid), severe + moderate (dotted line) and severe + moderate + mild infections (dashed line) are shown for age groups 0-19, 20-29, 30-39, 40-49, 50-59, 60-69, 70+, all ages. Daily reported COVID-19 infections are also shown (stars) (23).

#### Reduction in Contacts as per Model Fitting Results

The compliance, *κ*, to the public health mitigation phases are fit from the total incidence data (23). The value of *κ* is fit for each change in mitigation phase and to capture significant changes to compliance. The model is fit to total incidence data using a Least squares objective function that is minimized using the patternsearch algorithm implemented in MATLAB (39). The mitigation phases are detailed in supplemental section S8. Shown in Figure S2 are the combined effect of mitigation phase (black x’s) and the estimated value for *κ* (red pluses). Each marker (x and +) indicates the best-fit value for *κ*. The repeated markers with the same estimated value for *κ* during the vaccination program were fit together. Mitigation windows are indicated in Figure S2 with the green vertical lines and are detailed in Table S4.

**Figure S2:**
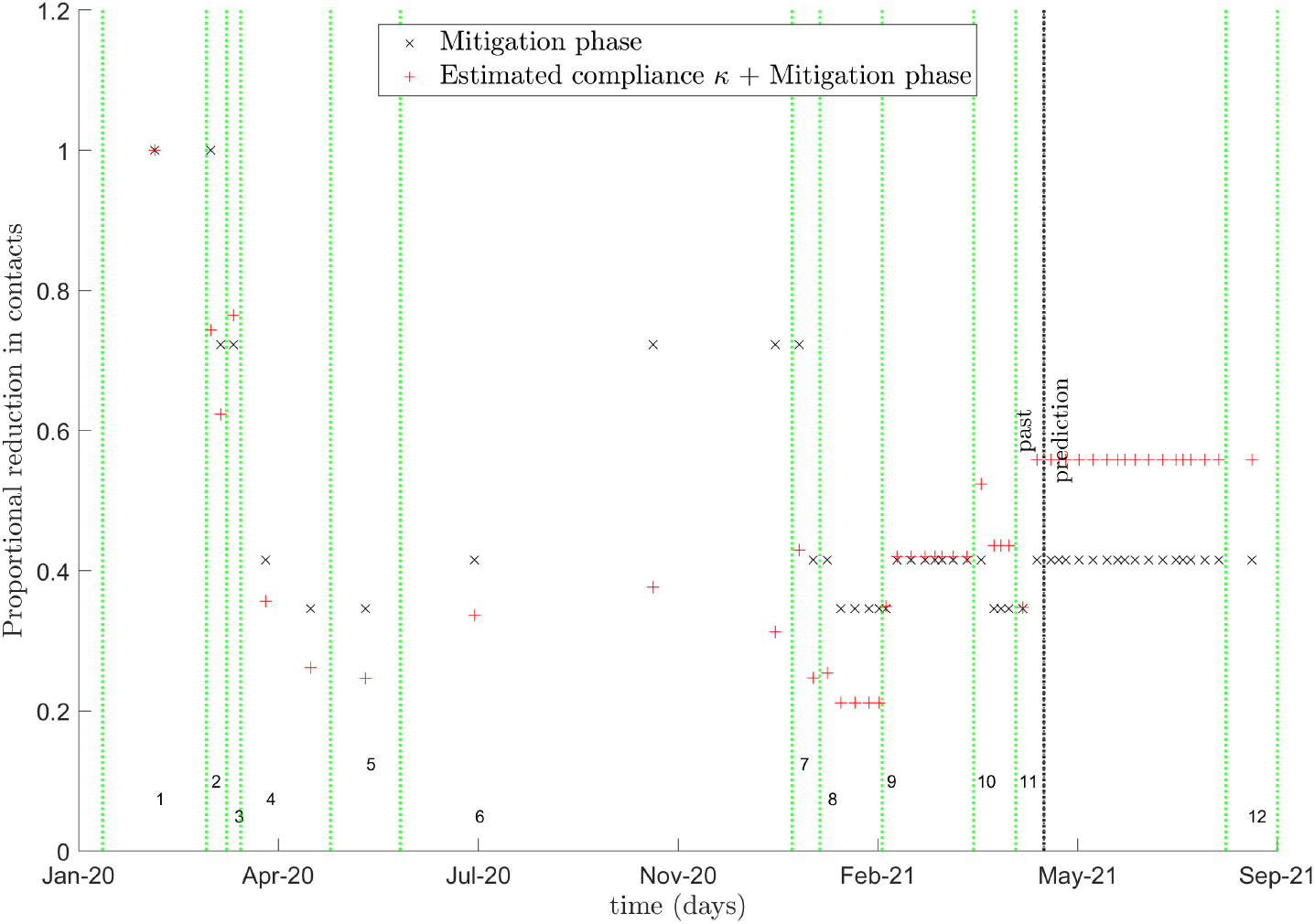
The reduction in contacts from mitigation phase (x) and the combined reduction in contacts, from mitigation phase and the estimated compliance, *κ*, to the public health mitigation (+). The x and plus markers are shown for each mitigation and vaccination window with the changes in mitigation phase indicated by the vertical dotted green lines. The mitigation windows are numbered at the bottom of the plot window. The start of the predicted mitigation phases and predicted combined effect of mitigation phase and *κ* are shown with the vertical dotted black, past/prediction, line.

#### Epidemiological effects of vaccination

As shown in Figures 3 and Supplementary Figure S3, all vaccination programs reduce the number of infections. A one-dose neutralizing vaccine (blue) has the best outcome, whereas a two-dose vaccine at half-coverage (orange) performs the worst. For all one- and two-dose vaccines, the best outcome for the entire population is realized when all adults 20+ years of age are vaccinated over the entire vaccination program (Scenario 4, dotted lines), but the best overall outcome in age groups 60+ years occurs under Scenario 1 (solid lines).

**Figure S3:**
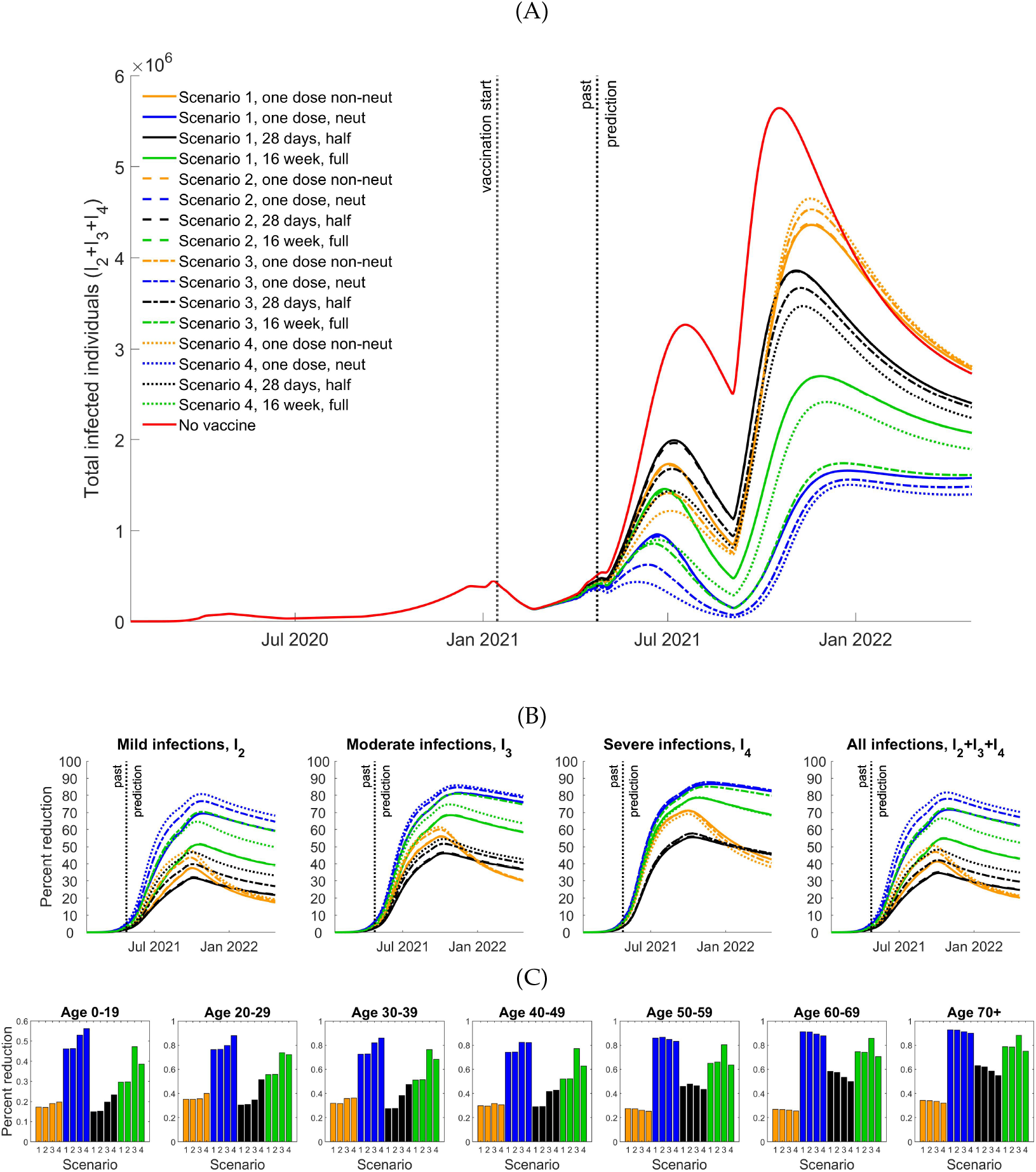
Effect of vaccination for Scenarios 1-4. (A) Infected populations without (red line) and with vaccination. (B) Percent reduction in *I*_2_, *I*_3_, *I*_4_, and *I*_2_ + *I*_3_ + *I*_4_ infections. Solid, dashed, dot-dashed, and dotted lines correspond to scenarios 1, 2, 3, and 4, respectively. (C) Percent reduction one year after vaccination program initiation Scenarios 1-4. All panels: one-dose non-neutralizing (orange), one-dose neutralizing (blue), two-dose half coverage (black), and two-dose full coverage (green) programs are shown.

#### Infections averted due to vaccination

*I*_4_ and total *I*_2_ + *I*_3_ + *I*_4_ infections averted due to all one- and two-dose vaccines under Scenarios 1-4 are shown in Figure S4. Once again, it is evident that maximum total infections averted result from Scenario 4, but that maximum total infections averted in ages 60+ years result from Scenario 1.

**Figure S4:**
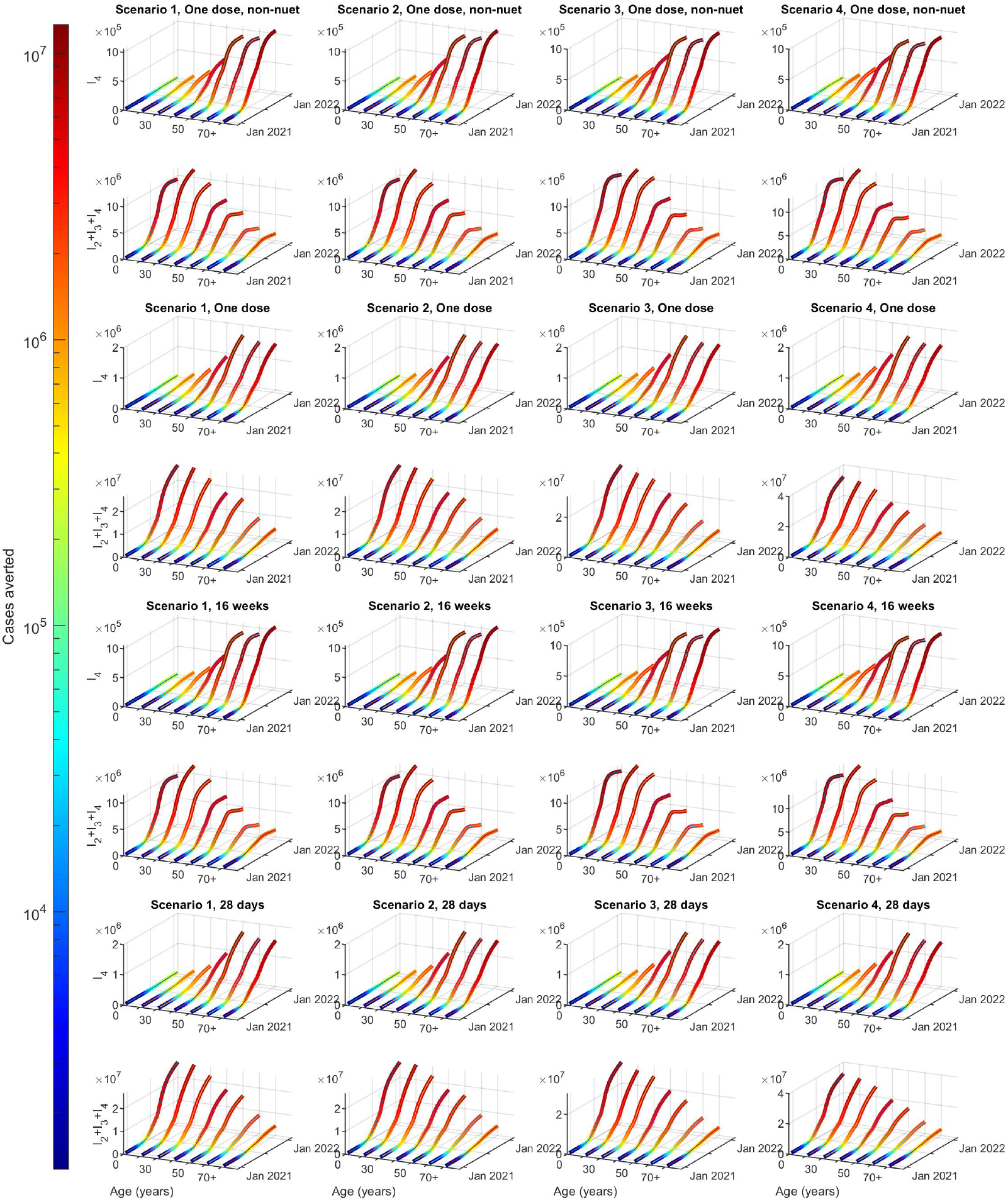
Daily infections averted. *I*_4_ and *I*_2_ +*I*_3_ +*I*_4_ infections averted under two-dose vaccination programs (doses separated by 16 weeks) with Scenarios 1 to 4 (columns). In paired rows, we plot cases averted in *I*_4_ (first row) and *I*_2_ + *I*_3_ + *I*_4_ (second row). Rows 1-2 are one dose non-neutralizing. Rows 3-4 are one dose, neutralizing. Rows 5-6 are two doses, half target coverage. Rows 7-8 are two doses, full target coverage. See Table 1 for vaccination program details.

#### Distribution of Daily Incidence

Figure S5 illustrates distributions of the daily incidence of *I*_4_ and *I*_2_ + *I*_3_ + *I*_4_ infections by age from September 2020 to January 2022, for Scenarios 1-4, under one- and two-dose vaccines (see Table 1).

**Figure S5:**
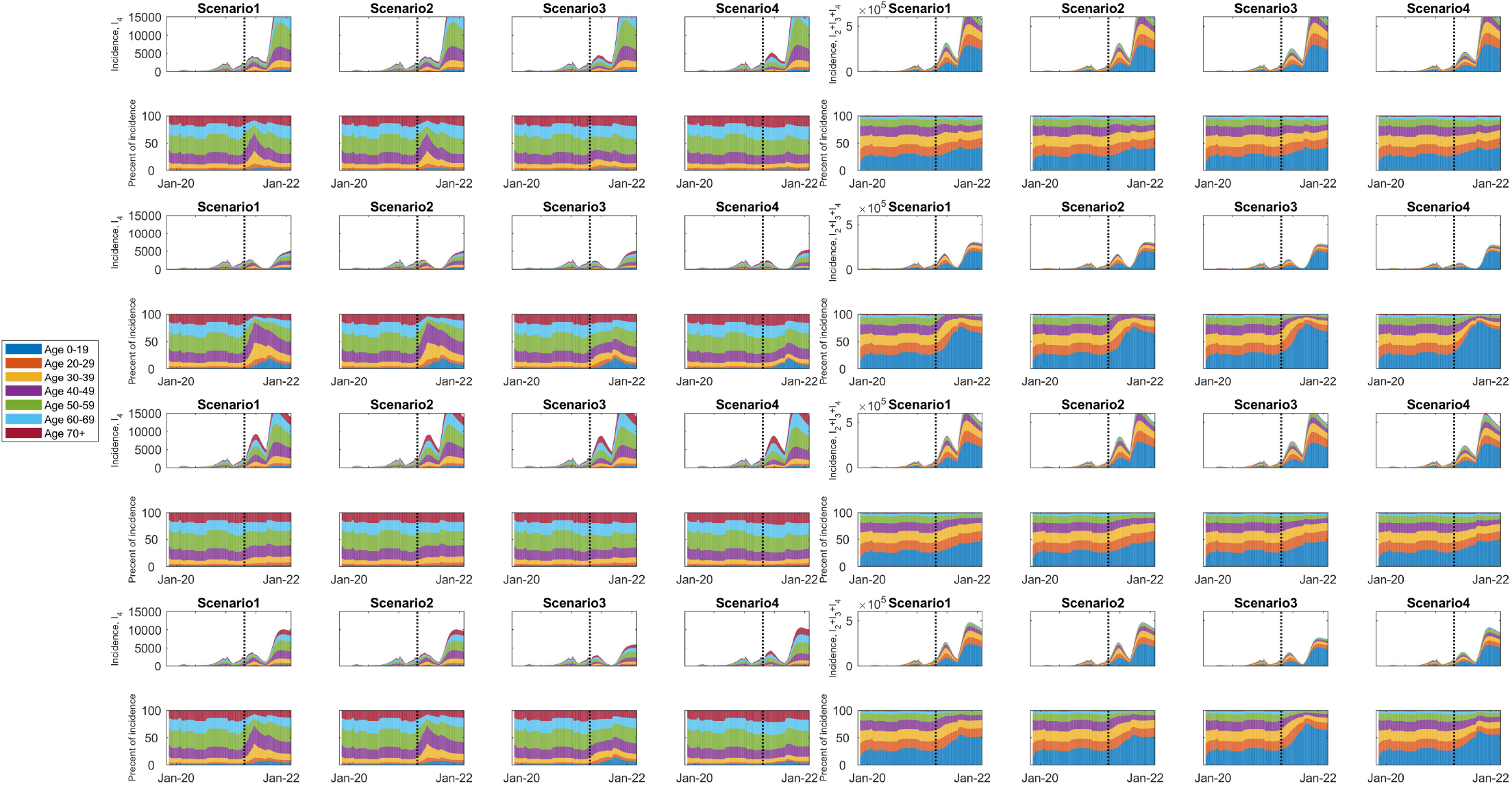
Age distribution of daily incidence with high target coverage. Columns are by scenario. In paired rows, we plot age distributions of the daily incidence of *I*_4_ and *I*_2_ + *I*_3_ + *I*_4_ infections (first row) and fractions of new infections in each age group (second row). Rows 1-2 are one dose non-neutralizing. Rows 3-4 are one dose, neutralizing. Rows 5-6 are two doses, half target coverage. Rows 7-8 are two doses, full target coverage. See Table 1 for vaccination program details.

#### Indirect Effect of Vaccination

Figure S6 illustrates the percent reduction in infections with mild *I*_2_ (first column), moderate *I*_3_ (middle column), and severe *I*_4_ (last column) symptoms given a two-dose vaccine, when 0.5% of the Canadian population is vaccinated each month for six months and all individuals vaccinated are from a single age group. Results are shown for 90 (top row), 180 (middle row) and 365 (bottom row) day periods after initiation of vaccination on December 15, 2020. The direct effect of vaccination is shown on the diagonals of each plot; all other elements reflect indirect effects.

**Figure S6:**
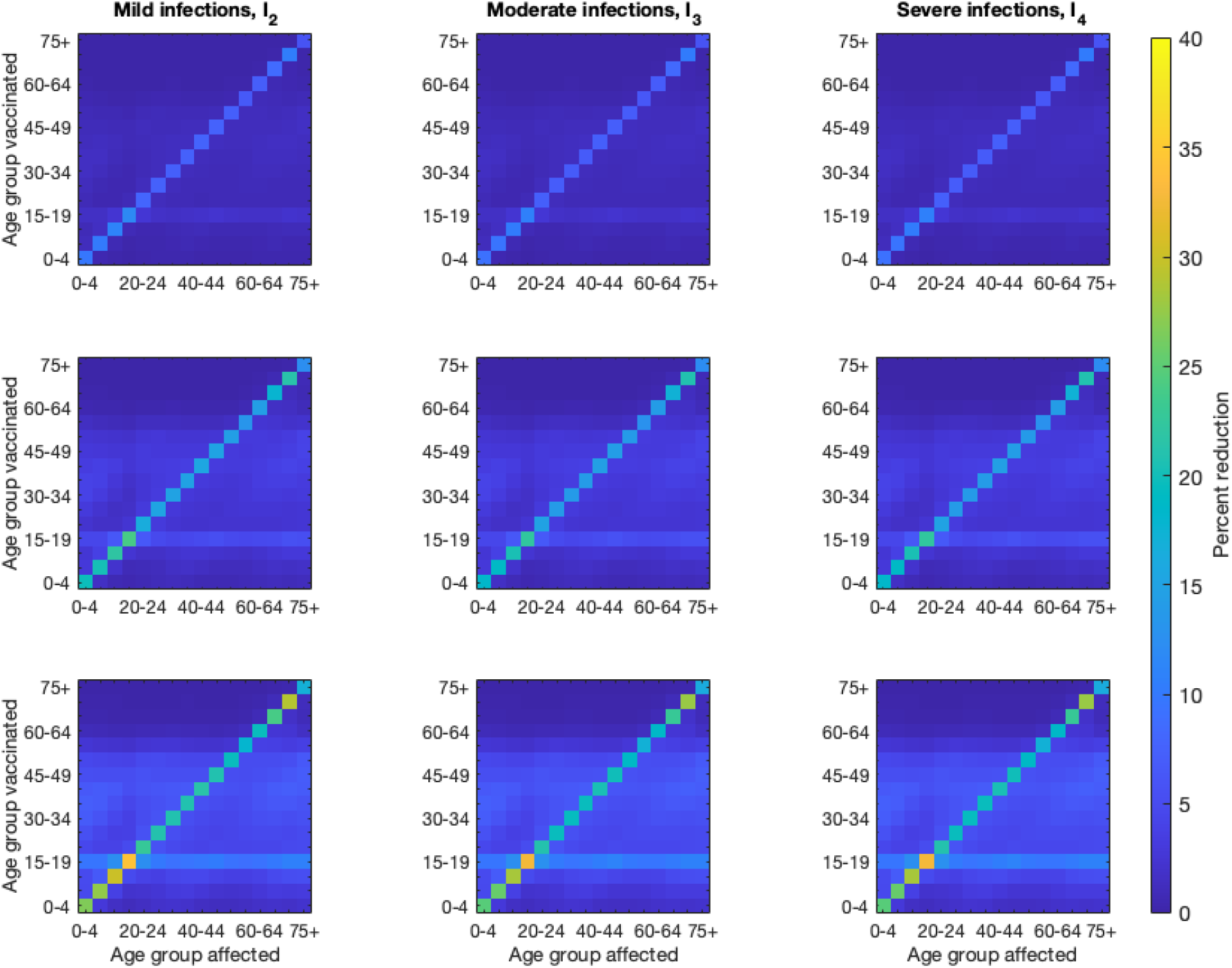
Indirect effects of vaccination. The percent reduction (color) in the number of mild (first column), moderate (middle column) and severe (third column) illnesses for each age group (x-axis), given administration of a two-dose vaccine (with 28 days separating doses, and two weeks until full realization of neutralizing immunity) to one age group (y-axis). The top row is after 90 days, the middle row is after 180 days, and the bottom row is after 365 days. Vaccination rates are calculated so that roughly 0.5% of the Canadian population is vaccinated each month for six months (up to 3% coverage over 6 months), but all vaccinated individuals are from a single age group (y-axis).

### Model

We track age, infection and immune status by modeling 16 age groups in five year intervals (ages 0-4, 5-9, …, 75+) and several susceptible (*S*), vaccinated (*V*) and infected (non-infectious *E*, infectious *I*) states. A schematic is provided in Figure S7 for a single age group. A description of the model is provided in the section entitled Model Description. Detailed descriptions of model parameters, including assumptions, are provided in the section entitled Parameters.

We distinguish four immune states (fully susceptible, somewhat immune, moderately immune, fully resistant to infection), and assume that individuals of higher immune status are less susceptible to infection than those of lower status. Immunity develops after infection, but the level depends on symptom severity. We assume that immunity wanes over time (Figure S7, black solid lines). Additionally, immunity is gained from vaccination with full resistance to infection occurring after two doses. We distinguish susceptible and vaccinated classes so that different assumptions regarding immunity gained after infection or vaccination could be tested. This also allows us to easily track population coverage under specified vaccination programs.

We use *S*_*i*_, 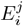, *I*_*i*_, and 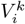 to denote the number of susceptible, exposed, infected and recovered individuals in each age group, where *i* (1 ≤ *I* ≤ 4) denotes immune status, 1 ≤ *j* ≤ 3 represents stages in the exposed class (gamma distributed), and *k* = 1, 2 denotes the number of doses of vaccine that individuals have received.

Here, for the *S* group, *i* = 1, …, 4, but for the *I* group, 2 ≤ *I* ≤ 4, where *i* is associated with the level of immunity that is acquired upon recovery from infection.

Susceptible individuals *S*_*i*_, *i* = 1, 2, 3, can be infected, with force of infection

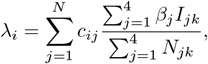

where *c*_*ij*_ denotes the proportion of contacts between age groups *i* and *j* and *β*_*j*_ is the probability of transmission for individuals with immune status *j*. We assume that infection also depends on the susceptible class, susceptibility index 0 ≤ *α*_*i*_ ≤ 1, and activity level *A*_*i*_. As more immune individuals are less susceptible, we assume that 1 = *α*_1_ ≤ *> α*_2_ ≤ *> α*_3_ ≤ *>* 0. Susceptible individuals *S*_*ij*_, upon infection, move to the mild, moderate, and severe symptom classes with probabilities 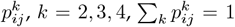. These probabilities are determined by the prevalence of no, one, and two or more co-morbidities within each age group. These probabilities are informed by (40).

Once infected, we assumed that individuals move through stages 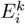, *i* = 2, 3, 4, *k* = 1, 2, 3 before they become infectious. We assumed that rates 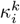 depend on the stage *k* and severity of infection *i*, allowing for different serial intervals and latent periods. Recovery and mortality rates depend on the severity of symptoms in each age class. We assume that for recovery *γ*_1_ *> γ*_2_ *> γ*_3_, and that for mortality *δ*_1_ *< δ*_2_ *< δ*_3_.

Susceptible individuals *S*_*i*_, *i* = 1, 2, 3, 4 can be vaccinated and move to 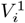. Then, upon a second dose, they move to 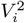, increasing their level of immunity. Vaccination rates 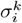, where *k* is the dose, are determined by coverage in each age group, and *ρ* represents the efficacy of the first dose of the vaccine. Vaccination rates 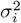 are determined by the number of days between the first and second doses of the vaccine, as well as the number of days needed to achieve full immunity from the second dose. In this report, we assume that individuals in 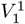 have similar immunity to those in susceptible class *S*_2_, and similarly, that 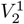 resembles *S*_3_. We assumed that all other vaccinated classes 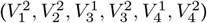 resemble *S*_4_ (i.e., are entirely resistant to infection).

Detailed descriptions of the model variables, parameters and equations are provided in sections Model Equations and Parameters, including assumptions and references.

**Figure S7:**
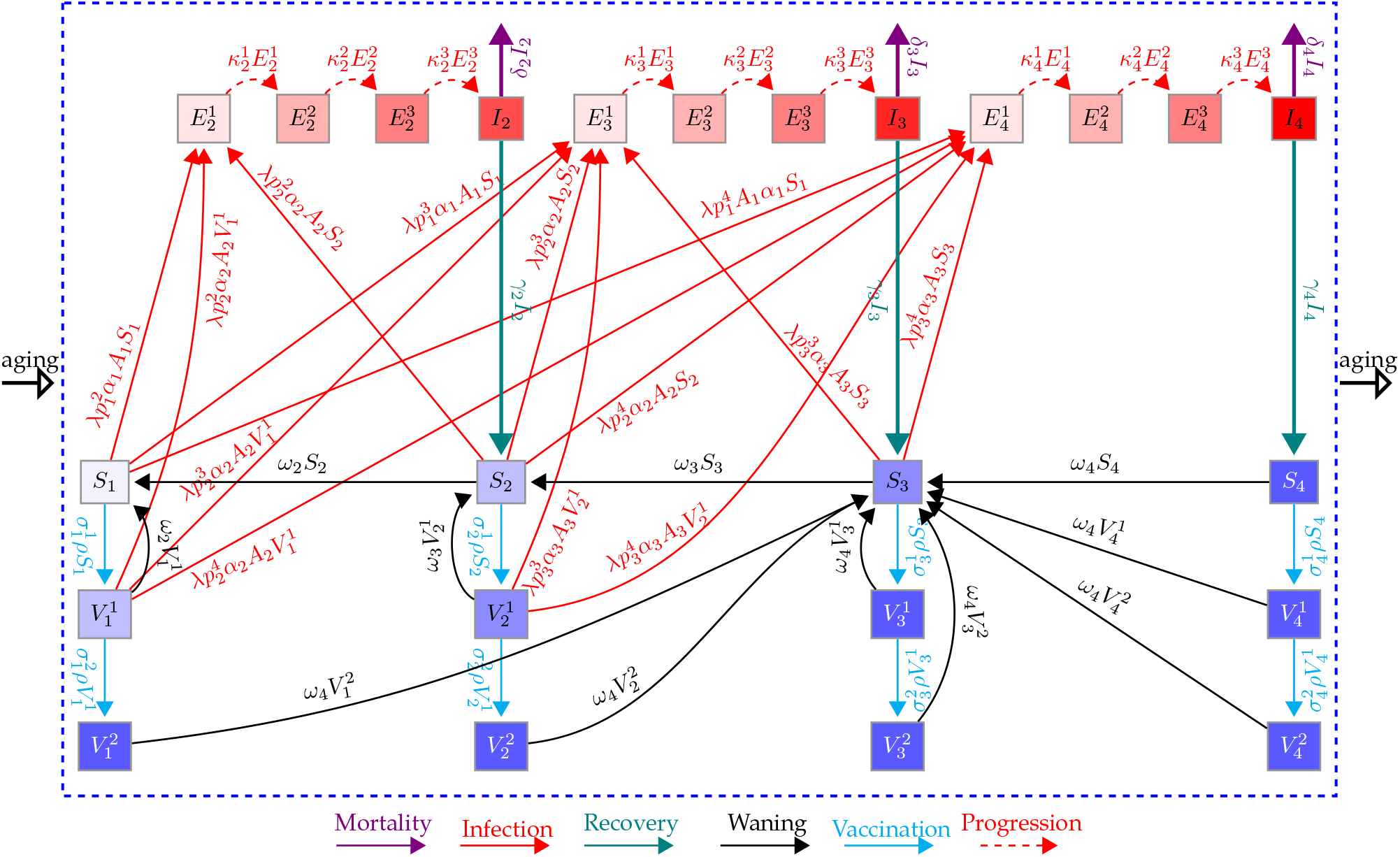
Schematic of the model for one age group. Here, *S*_1_, *S*_2_, *S*_3_, and *S*_4_ (purple shaded boxes) represent susceptible individuals who are immunologically naive, have some immunity, and are moderately and fully immune, respectively. *I*_2_, *I*_3_, and *I*_4_ (red boxes) represent infected individuals with mild, moderate and severe symptoms, respectively, who will develop some, moderate, and full immunity upon recovery (teal solid line), respectively. 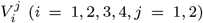 represent vaccinated individuals from the *S*_*i*_ classes (*i* = 1, 2, 3, 4) after *j* = 1, 2 doses of a two-dose vaccine. 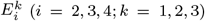 represent exposed individuals (infected, asymptomatic, not infectious) with progressive stages *k* = 1, 2, 3, that will eventually experience mild *I*_2_, moderate *I*_3_, and severe *I*_4_ symptoms. Susceptible and vaccinated individuals can be infected and move to the exposed classes (red lines). Similarly, susceptible and vaccinated classes at the same location on the immunity continuum have the same characteristics. Immunity gained from infection and vaccination can wane over time (black lines).

#### Model Equations

Our model tracks age, infection and immune status. Susceptible individuals of status *i* and age *k* are denoted by *S*_*ik*_; similarly, infectious individuals are denoted by *I*_*ik*_. Infected, but not-yet-infectious, individuals of immune status *i*, age *k* and stage *j* are denoted by 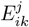. Vaccinated individuals of immune status *i*, age *k* and dose *k* are denoted by 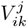. Parameter descriptions are found in Table S2. The system of ODEs for age group *k* is given by the following set of equations:

Susceptible compartments:

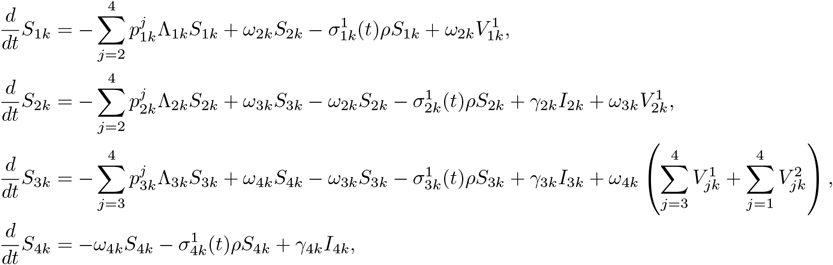

Vaccinated compartments:

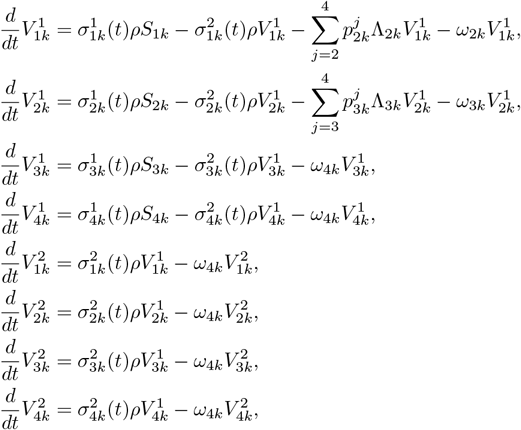

Infected compartments:

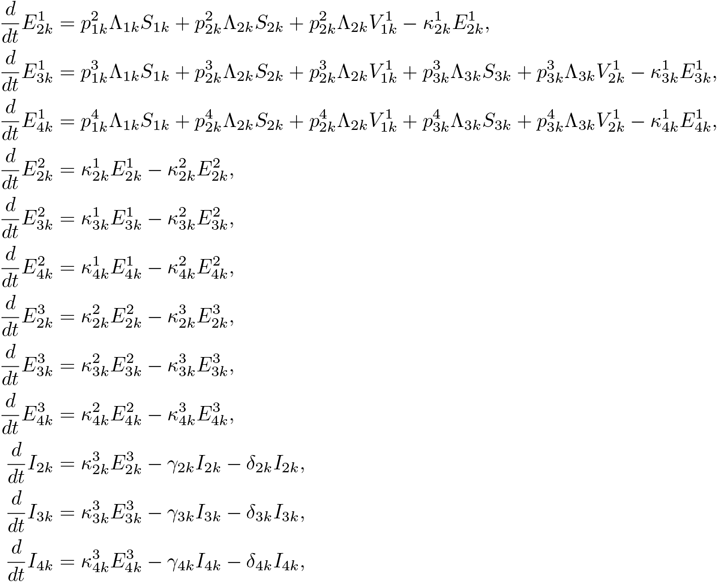

where for 1 *≤ k ≤ N*,

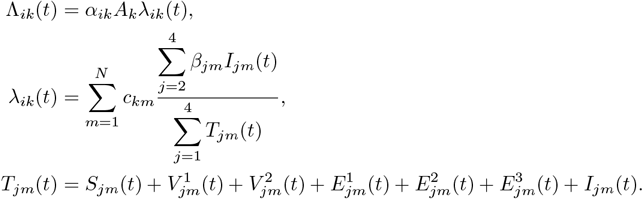

### Parameters

**Table S2:** Parameter definitions for the ODE model.

| Parameter | Definition |
| --- | --- |
| $\alpha_{im}$ | susceptibility of individuals from $S_{im}$ ($i$ immunity status, $m$ age group) |
| $\beta_{jm}$ | infectivity of individuals from $I_{jm}$ ($j$ immunity status, $m$ age group) |
| $\gamma_{jm}$ | recovery rate of individuals from $I_{jm}$ ($j$ immunity status, $m$ age group) |
| $\kappa_{jm}^k$ | rates of progress through the pre-infectious period ($i$ immunity status, $m$ age group, $k$ stage) |
| $\delta_{jm}$ | disease-induced mortality rate of infected individuals from $I_{jm}$ ($j$ immunity status, $m$ age group) |
| $\omega_{im}$ | waning rate of immunity of individuals from $S_{im}$ ($i$ immunity status, $m$ age group) |
| $p_{im}^j$ | proportion of $S_i$ going to $I_j$ upon infection with $i = 1, 2, 3$ and $j = 2, 3, 4$ ($i, j$ immunity status, $m$ age group) |
| $\rho_{im}$ | vaccine efficacy ($i$ immunity status, $m$ age group) |
| $\sigma_{jm}^k$ | vaccination rate for first $k = 1$ and second $k = 2$ dose ($j$ immunity status, $m$ age group) |
| $A_n$ | per capita contact rate of individuals in age group $n$ |
| $c_{an}$ | mixing between individuals in age group $a$ and age group $n$ |

#### Disease Parameters

Susceptibility is assumed to decrease with increasing immunity, but not depend on age. Thus, *α*_1*n*_ = 1, 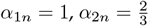, and 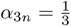 for any age group *n*. Infectivity is assumed to vary by severity of disease and was chosen to produce a basic reproduction number *R*_0_ = 2.6. By immunity status, we assume *β*_3*n*_ = *β* = 0.08, *β*_2*n*_ = 0.5*β*, and *β*_4*n*_ = 0.1*β* for any age group *n*. People with milder symptoms are expected to have lower viral loads and, thus, lower infectivity. Simultaneously, more severe disease outcomes are expected to induce behavioral changes, such as limiting mobility, that lower infectivity.

The recovery rate is assumed to depend on symptom severity, with milder disease associated with shorter infectious periods. Thus, *γ*_1*n*_ *< γ*_2*n*_ *< γ*_3*n*_, with 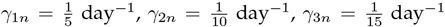 for any age group *n*. We assume that disease-induced mortality only occurs among those with the most severe symptoms, *I*_4_. Thus, *δ*_2*n*_ = 0, *δ*_3*n*_ = 0, *δ*_4*n*_ = *δ* = 0.0001 for any age group *n*.

Every susceptible class can become infected and recover with equal or greater immunity. The proportion of any susceptible class experiencing more severe symptoms increases with age; i.e., 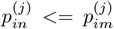 for *n < m*. Here, we assume that age is a proxy for co-morbidity. Specific values were determined from (40).Values are found in Table S3.

**Table S3:** Proportion of susceptible individuals from *S*_*in*_ going to *I*_*j*_ infectious class, given by 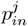.

| Age group | $p_{in}^{(2)}$ | $p_{in}^{(3)}$ | $p_{in}^{(4)}$ |
| --- | --- | --- | --- |
| 0-4 | 0.979187625 | 0.019532353 | 0.001280022 |
| 5-9 | 0.970340674 | 0.02795418 | 0.001705146 |
| 10-14 | 0.971354725 | 0.026962603 | 0.001682673 |
| 15-19 | 0.963790465 | 0.033836424 | 0.002373111 |
| 20-24 | 0.94736408 | 0.048618385 | 0.004017534 |
| 25-29 | 0.923743993 | 0.069289774 | 0.006966234 |
| 30-34 | 0.897203802 | 0.09151371 | 0.011282488 |
| 35-39 | 0.865605311 | 0.116760519 | 0.01763417 |
| 40-44 | 0.806984827 | 0.162542801 | 0.030472373 |
| 45-49 | 0.756587998 | 0.197861718 | 0.045550284 |
| 50-54 | 0.690245134 | 0.241501371 | 0.068253495 |
| 55-59 | 0.600190415 | 0.296345592 | 0.103463993 |
| 60-64 | 0.503245046 | 0.346804634 | 0.14995032 |
| 65-69 | 0.409505408 | 0.383960071 | 0.206534521 |
| 70-74 | 0.324664092 | 0.404030163 | 0.271305745 |
| 75+ | 0.215150255 | 0.373779207 | 0.411070537 |

We assume that immunity lasts on average one year between successive stages and is independent of age. Thus, the waning of immunity occurs at rate 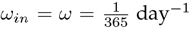 for *i* = 1, 2, 3 and age group *n*. For vaccinated classes, we assume that immunity in 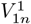 wanes to *S*_1*n*_, that in 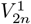 wanes to *S*_2*n*_, and that of all other vaccinated classes wanes to *S*_3*n*_.

#### Vaccination Parameters

The first dose vaccination rate is determined by the coverage detailed in the NACI document (24). Monthly coverage, *y*_*in*_, is scaled to a daily rate, 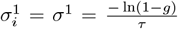 where *g* accounts for the portion of the population in the current vaccine-eligible age groups, *I*_4*n*_, 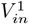, and 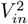, that we assume are not allowed to be vaccinated, either due to having severe symptoms or prior vaccination. Our calculation assumes that each month has, on average, 30 days. Vaccine efficacy for each dose is assumed to be 0.9. Individuals receive a second dose 28 days or 16 weeks (112 days) after the first and acquire immunity after an additional 14 days. Thus, the vaccination rate of the second dose is 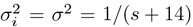, where *s* = 28 or 112 days.

#### Demographic Parameters

Given the short period considered, we assume the absence of natality, natural mortality and aging. We focus on the importance of contacts among age groups. We assume that the mixing matrix and associated activities only depend on age and are taken from (41). The base matrix provides the number of contacts between ages at school (S), work (W), home (H) and in other (O) places. We also determine matrices for different stages of mitigation and relaxation: strict mitigation (phase 0); moderate mitigation (phase 1); moderate relaxation (phase 2); and increased relaxation (phase 3). These contact matrices were scaled by a compliance index, *κ* (see Figure S2). These contact matrices and compliance values were chosen in accordance with Canadian policy.

Phase 0 (strict mitigation) is characterized by large reductions in all contacts compared to baseline. There is a 95% reduction of school contacts for all age groups. For individuals below age 65, there is a 75% reduction in work contacts and a 90% reduction in other contacts. For ages above 65, work and other contacts are reduced by 95%. Phase 1 (moderate mitigation) retains the same reduction in school contacts as phase 0, but eases reductions in work and other contacts slightly. For ages below 65, there is a 70% reduction compared to baseline for work contacts, while the reduction remains the same as phase 0 for older ages. For all ages, other contacts are reduced to 25% of baseline, a more than doubling from phase 0. Phase 2 (moderate relaxation) is characterized by a rebound in contacts in all areas. School contacts are only reduced by 15% compared to baseline for age groups below age 65. For ages below 65, work contacts are reduced by 35% and other contacts by 40% compared to baseline. Older ages remain with a 95% reduction in work contacts, but only a 65% reduction in other contacts. Phase 3 (increased relaxation) nearly returns to normal contacts. School contacts for those below 65 are at a 5% reduction. For all ages, work contacts and other contacts are reduced by 25%.

Perturbations of the contact matrices are implemented to reflect public health mitigation strategies over time. A summary of the perturbations is given in Table S4.

**Figure S8:**
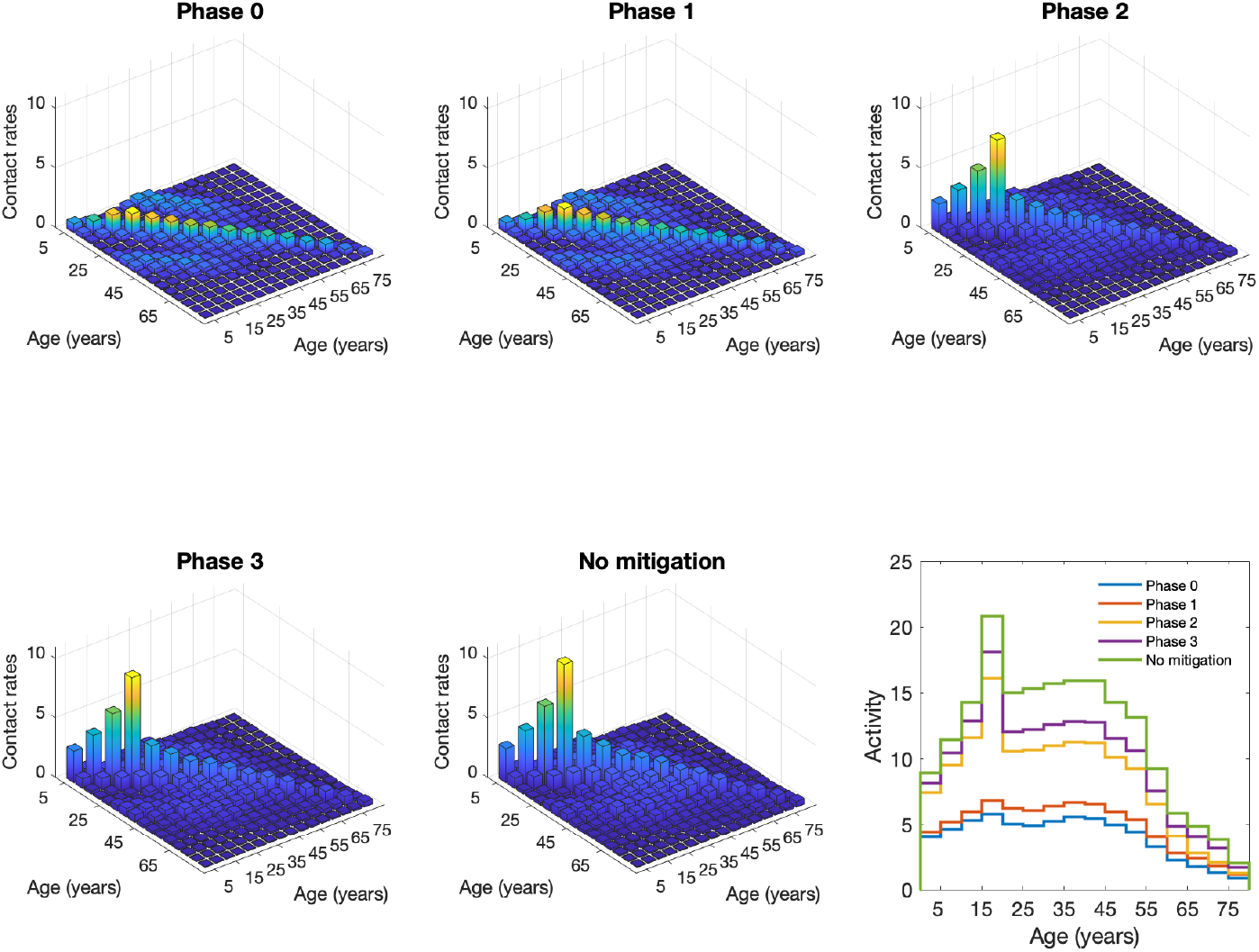
Age-specific and marginal contact rates under different phases of mitigation. Bar plots show contact rates between age groups under different mitigation strategies, with compliance (*κ*) assumed to be 100%. Taller bars, by subplot, have lighter colors. All bar plots have identical scaling for comparison. The total activity is shown in the line plot (bottom right).

#### Age Distribution of Healthcare and Personal Support Workers

There are approximately 1.45 million healthcare providers in Canada (25). Additionally, there are at least 100,000 personal support workers (PSW) (27). Our framework does not explicitly model these populations. Considering the prioritization of these groups in vaccination campaigns, we must distribute them into modeled age groups. A recent report (25) shows that approximately 15%, 73%, and 12% of healthcare workers are aged *<*30, 30-59, and 60+ years, respectively. An older, but more detailed report shows that [14%, 12%, 11%, 11%, 12%, 13%, 12%, 13%] of all registered nurses in year 2015 had ages [*<*30, 30-34, 35-39, 40-44, 45-49, 50-54, 55-59, 60+], respectively. A report on long-term care staffing (27) states that 50% of PSWs working in the healthcare sector lie between the ages of 35 and 54, and that 25% of the remaining PSWs are 55+ years of age. In the older report (28), [0.3%, 7%, 18.7%, 29.2%, 32.5%, 12.3%] of PSWs were aged [*<*20, 20-29, 30-39, 40-49, 50-59, 60+] years. Given that the combined distributions of healthcare and personal support workers is approximately uniform, we assume that there is an even distribution of these priority workers in age groups 20-64.

**Table S4:** Contact Perturbation Summary

| Contact mitigation phase description (% reduction in contacts) |  |  |  |
| --- | --- | --- | --- |
| Phase | School Contacts | Other Contacts | Work Contacts |
| 0 | 95% | 90% under 65, 95% over 65 | 75% under 65, 95% over |
| 1 | 95% | 75% | 70% under 65, 95% over |
| 2 | 15% under 20, 25% over | 75% | 70% under 65, 95% over |
| 3 | 15% under 20, 25% over | 75% | 35% under 65, 95% over |
| no mitigation | 0% | 0% | 0% |

| Contact mitigation phase |  |  |
| --- | --- | --- |
| Start | End | Phase |
| 2020-02-05 | 2020-03-18 | no mitigation |
| 2020-03-18 | 2020-03-31 | 3 |
| 2020-03-31 | 2020-04-25 | 1 |
| 2020-04-25 | 2020-06-19 | 0 |
| 2020-06-19 | 2020-09-01 | 1 |
| 2020-09-01 | 2021-01-08 | 3 |
| 2021-01-08 | 2021-01-22 | 1 |
| 2021-01-22 | 2021-02-19 | 0 |
| 2021-02-19 | 2021-04-09 | 1 |
| 2021-04-09 | 2021-05-14 | 0 |
| 2021-05-14 | 2021-09-01 | 1 |
| 2021-09-01 | 2021-12-31 | 3 |
| 2021-09-01 | 2021-12-31 | 3 |

## Notes

### Competing Interest Statement

The authors have declared no competing interest.

### Author Declarations

We followed all relevant ethical guidelines and a formal ethics approval was not needed for our modelling study.

